# Long-Term Clinical Performance of the Tunnel Technique with Subepithelial Connective Tissue Grafting: A 16-Year Retrospective Cohort Study

**DOI:** 10.64898/2026.08.17.26360439

**Authors:** Jennifer Schmücker, Emily Speer, Marco Alexander Vukovic, Wolf-D. Grimm

**Author notes:** Corresponding author: Wolf-D. Grimm, Univ.-Prof. Dr. Dr.Senior Professor, Periodontology, University of Witten/Herdecke, Germany, Practice Team Hasslinghausen, Center of Excellence for Periodontology and Implantology of the DGParo from 2019 to 2025, Sprockhoevel, Germany.

## Abstract

**Background:** Subepithelial connective tissue grafting remains a reference treatment for predictable root coverage. Although short- and medium-term outcomes of tunnel-based procedures are well documented, evidence regarding stability beyond 10 years remains limited. This study evaluated the long-term clinical performance of a minimally invasive tunnel technique combined with subepithelial connective tissue grafting (SCTG) under routine clinical conditions.

**Methods:** This retrospective longitudinal cohort study included 74 patients (57 women and 17 men) contributing 710 gingival recession sites treated between 2009 and 2025. All sites were treated with a tunnel approach and SCTG, with enamel matrix derivative (EMD) used in selected cases. The mean follow-up was 6.0 ± 4.0 years, with a maximum observation period of 16 years. The primary outcome was recession depth reduction. Secondary outcomes included complete root coverage (CRC), mean root coverage, and long-term marginal stability. Clinically relevant relapse was defined as a ≥1 mm increase in recession after initial healing.

**Results:** Mean recession reduction was 2.72 mm. Complete root coverage was achieved at 83.4% of treated sites. At the final available follow-up, no treated site showed a clinically relevant relapse of ≥1 mm after initial healing, and no site deteriorated beyond its baseline recession level. Treatment effects were observed across anterior and posterior regions.

**Conclusions:** Within the limitations of a retrospective cohort design, tunnel surgery combined with SCTG was associated with high root-coverage predictability and durable marginal soft-tissue stability for observation periods extending to 16 years. These real-world data support phenotype-enhancing, minimally invasive soft-tissue augmentation as a durable therapeutic strategy for localized and multiple gingival recessions.

## 1. Introduction

Periodontal plastic surgery has progressively moved toward minimally invasive microsurgical concepts that prioritize preservation of the vascular supply, atraumatic tissue handling, stable wound closure, and modification of the periodontal soft-tissue phenotype. In the management of gingival recession, autogenous connective tissue grafting remains the most extensively documented approach for enhancing root coverage and increasing soft-tissue thickness.

The tunnel technique was developed to achieve coronal mobilization of the marginal tissues while preserving papillary integrity and avoiding vertical releasing incisions. By maintaining a broad vascular bed around the graft, the technique is intended to facilitate revascularization, wound stability, and harmonious soft-tissue integration. Randomized trials and systematic reviews have reported favorable complete root coverage rates for tunnel-based procedures, particularly when combined with an autogenous connective tissue graft.

Beyond initial root coverage, long-term stability is increasingly recognized as a clinically important endpoint. Longitudinal investigations indicate that gains in soft-tissue thickness and phenotype modification may influence the maintenance of the gingival margin over time. Nevertheless, most controlled studies have observation periods of only months to a few years, whereas large real-world cohorts followed for more than a decade remain uncommon.

The present retrospective longitudinal cohort study therefore evaluated the clinical performance of the tunnel technique combined with SCTG, with or without adjunctive EMD, in routine periodontal practice. The primary objective was to quantify recession reduction.

Secondary objectives were to determine complete root coverage and to assess marginal stability during follow-up extending to 16 years. The working hypothesis was that minimally invasive tunnel surgery combined with phenotype enhancement by SCTG would be associated with sustained root coverage and a low frequency of clinically relevant relapse.

## 2. Materials and Methods

### 2.1 Study design and setting

This investigation was designed as a retrospective longitudinal cohort study of patients treated for gingival recession in a private periodontal referral setting. Surgical procedures were performed between 2009 and 2025 by one experienced periodontist using a standardized microsurgical tunnel approach. Clinical records available through the final observation period were reviewed to identify eligible sites and the last documented follow-up for each site.

**Figure 1.**
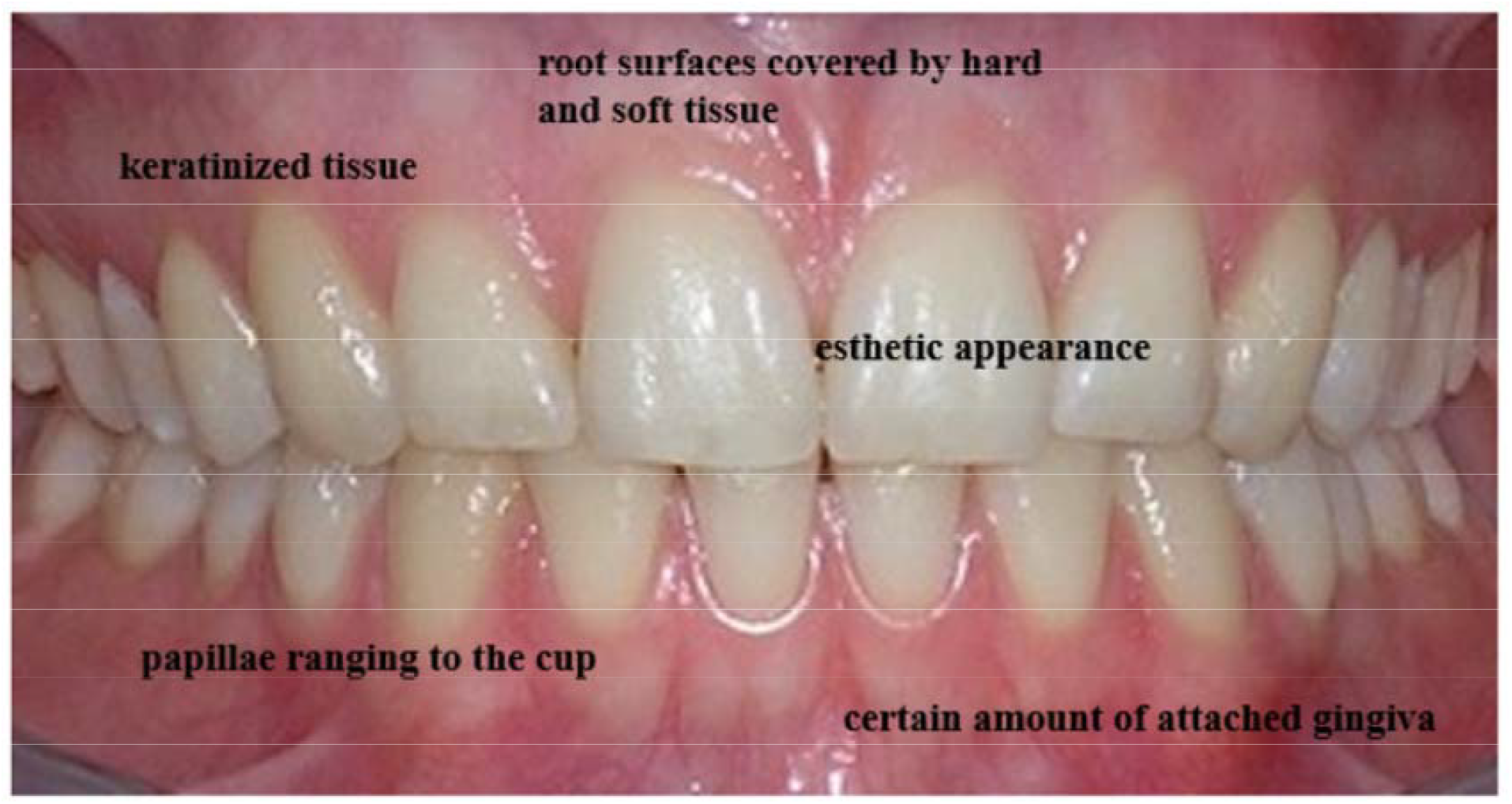
Clinical characteristics of an aesthetically favorable gingival architecture, illustrating adequate keratinized/attached tissue, papillary fill, and soft-tissue coverage.

### 2.2 Study population

The final cohort comprised 74 patients (57 women and 17 men) and 710 treated recession sites. Teeth from multiple regions of the maxilla and mandible were represented. Each treated tooth was considered a site-level analytical unit. Mean clinical follow-up was 6.0 ± 4.0 years; the longest documented observation period was 16 years.

### 2.3 Eligibility criteria

Sites were eligible when they presented with gingival recession of at least 3 mm, were classified as Miller Class I-III, and had complete baseline and follow-up documentation. Where indicated, non-surgical periodontal therapy was completed before surgery. At the time of surgical treatment, full-mouth plaque scores were <20%, full-mouth bleeding scores were <15%, and probing depths at treated sites were ≤4 mm.

Exclusion criteria were uncontrolled systemic disease, clinically relevant immunosuppression, pregnancy or lactation, previous head-and-neck radiation therapy, or incomplete clinical documentation.

### 2.4 Surgical protocol

All procedures followed a minimally invasive microsurgical protocol. After local anesthesia, intrasulcular incisions were performed without vertical releasing incisions. A tunnel was prepared around the recession defects, with full-thickness elevation in the coronal portion and a transition to split-thickness preparation apically, permitting tension-free coronal mobilization beyond the mucogingival junction. Interdental papillae were preserved.

**Figure 2.**
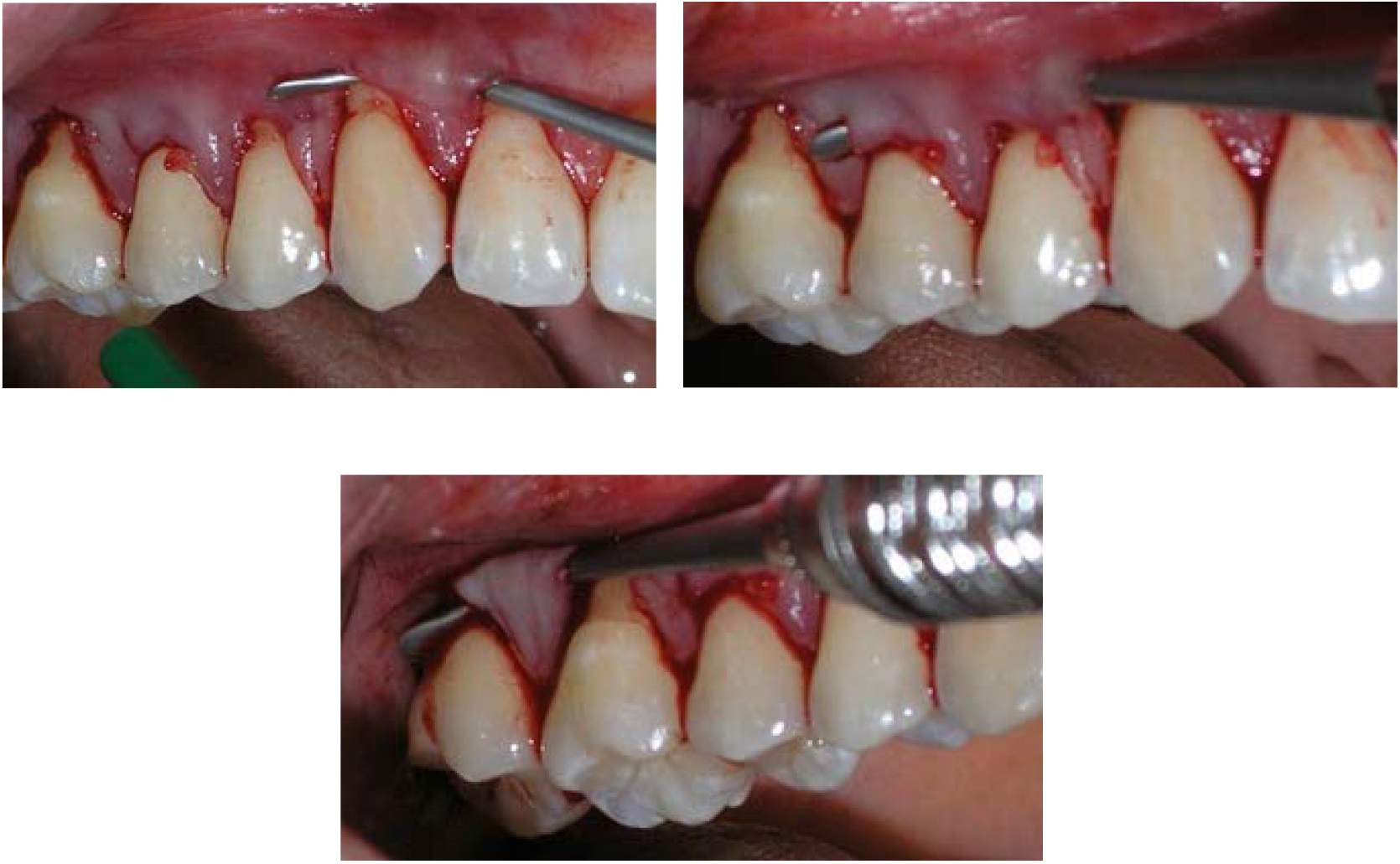
Incision-free tunnel preparation used for minimally invasive root-coverage surgery.

Subepithelial connective tissue grafts were harvested from the palate using a single-incision approach intended to minimize donor-site morbidity. The graft was introduced into the prepared tunnel and stabilized with sling sutures. In selected cases, EMD was applied as an adjunct according to the clinical indication and the treatment protocol in use at the time.

**Figure 3a.**
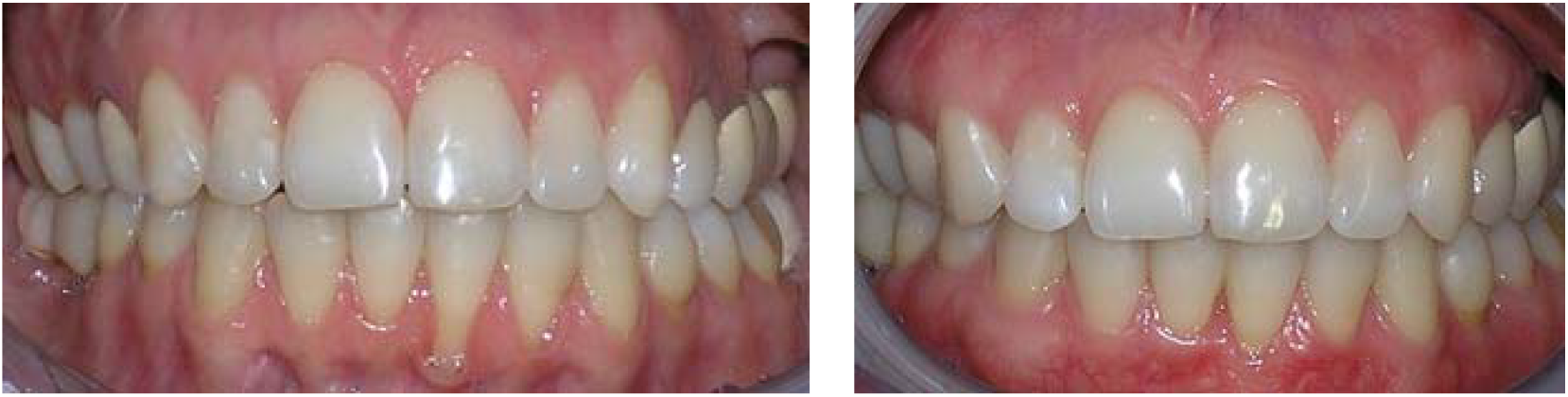
Representative clinical case at baseline and after 12 years.

**Figure 3b.**
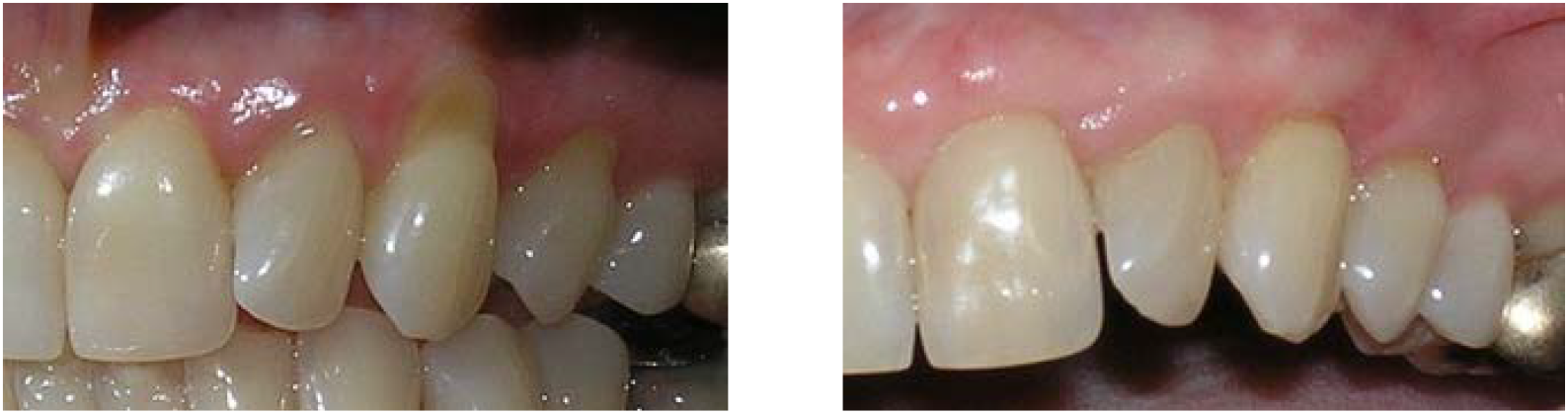
Representative clinical case at baseline and after 15 years.

**Figure 4.**
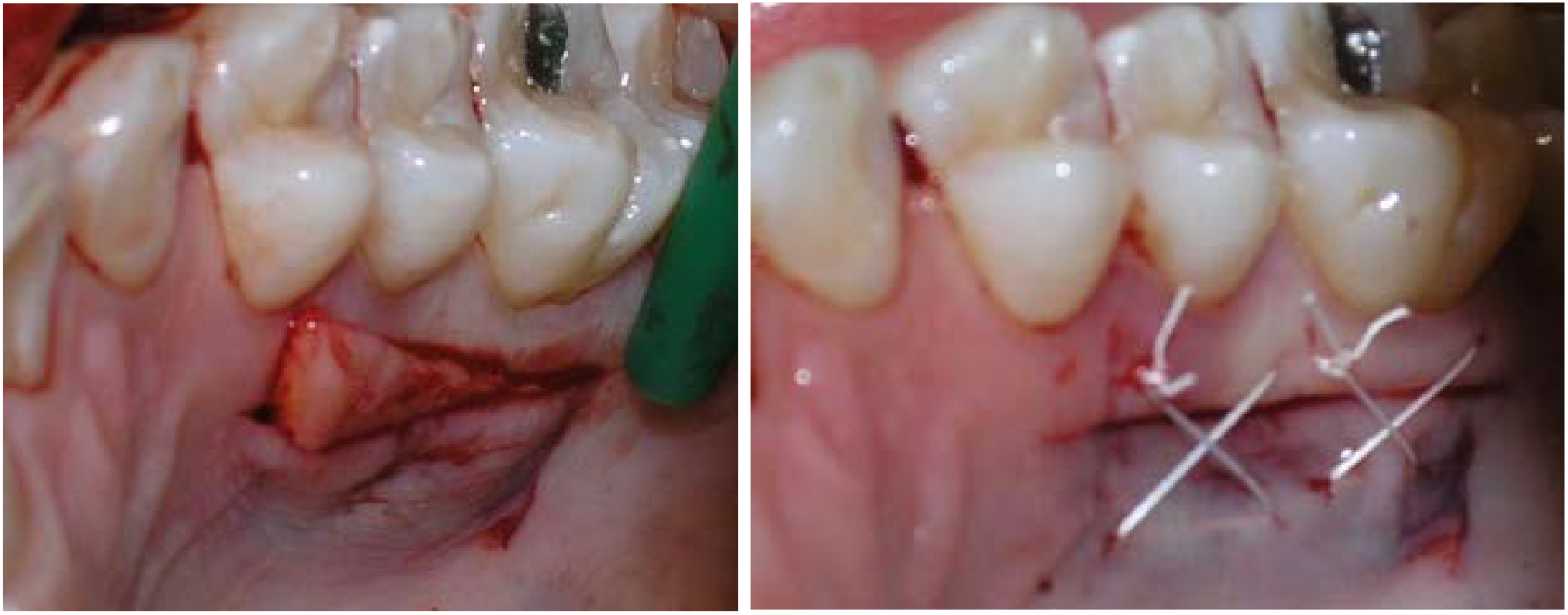
Harvesting of a subepithelial connective tissue graft using the single-incision technique.

Postoperative care included chlorhexidine rinsing and standardized oral-hygiene instructions. Patients subsequently entered supportive periodontal therapy, with recall intervals generally ranging from 3 to 12 months according to individual periodontal risk.

### 2.5 Outcome measures

The primary outcome was recession depth (RD), expressed in millimeters, and the corresponding change from baseline to the last available follow-up. Complete root coverage (CRC) was defined as a final RD of 0 mm. Mean root coverage (MRC) was considered as a secondary root-coverage measure when the underlying clinical measurements were available. Long-term stability was assessed from the longitudinal clinical documentation. A clinically relevant relapse was prespecified as an increase in recession of ≥1 mm after initial healing.

### 2.6 Statistical analysis

Descriptive statistics were summarized as means and standard deviations for continuous variables and as frequencies and percentages for categorical outcomes. Baseline and follow-up measurements were compared using paired parametric or non-parametric procedures, as appropriate to the distribution of the data. Because multiple treated sites could originate from the same patient, within-patient clustering was addressed using a repeated-measures framework. Statistical significance was defined at α = 0.05. The present preprint reports the clinically documented cohort-level outcomes available in the source dataset.

## 3. Results

### 3.1 Cohort and follow-up

Seventy-four patients contributed 710 recession sites to the final analysis. All included sites had sufficient baseline and follow-up documentation for longitudinal evaluation. The mean follow-up was 6.0 ± 4.0 years, and the maximum observation period was 16 years.

**Figure 5.**
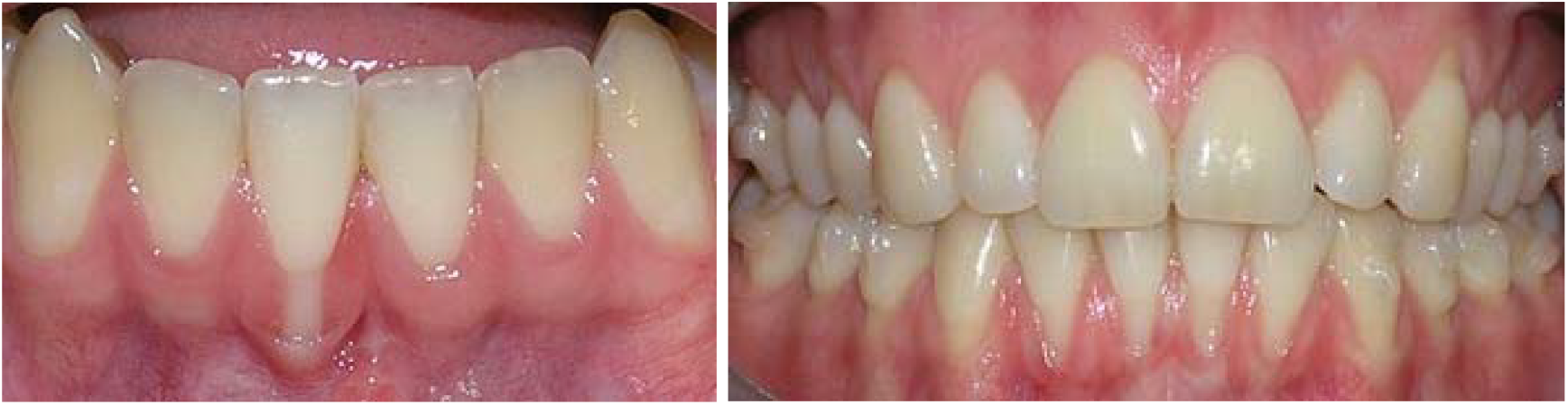
Representative root-coverage cases from the clinical cohort.

### 3.2 Recession reduction and complete root coverage

Treatment was associated with a marked reduction in gingival recession. Mean recession reduction across the cohort was 2.72 mm per treated site. Complete root coverage, defined as a final recession depth of 0 mm, was documented at 83.4% of sites. Residual recession in the remaining sites was limited, and no site showed deterioration beyond its pretreatment recession level at the final recorded follow-up.

**Figure 6.**
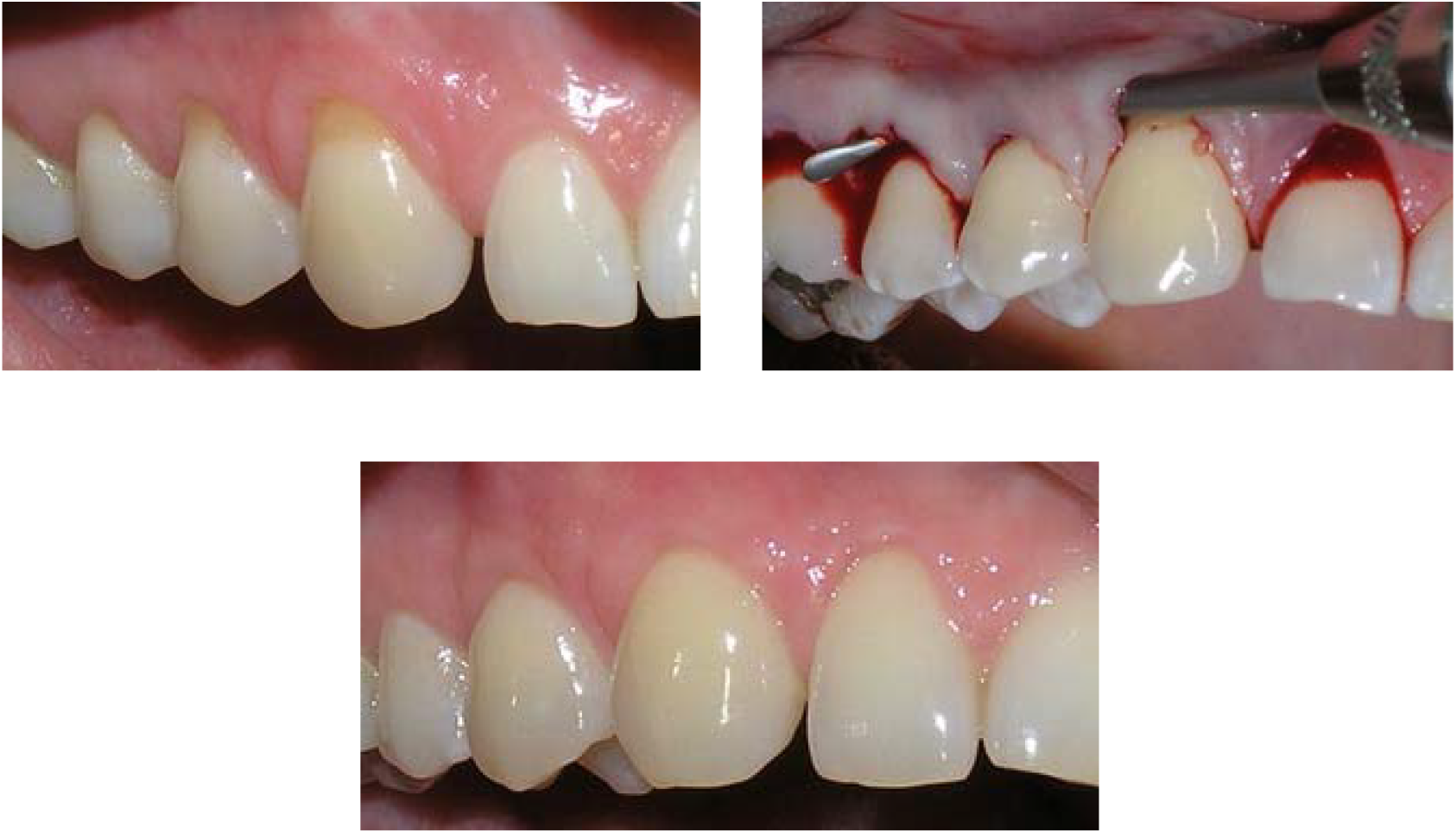
Representative case showing baseline recession, tunnel treatment, and the clinical result after 15 years.

**Figure 7.**
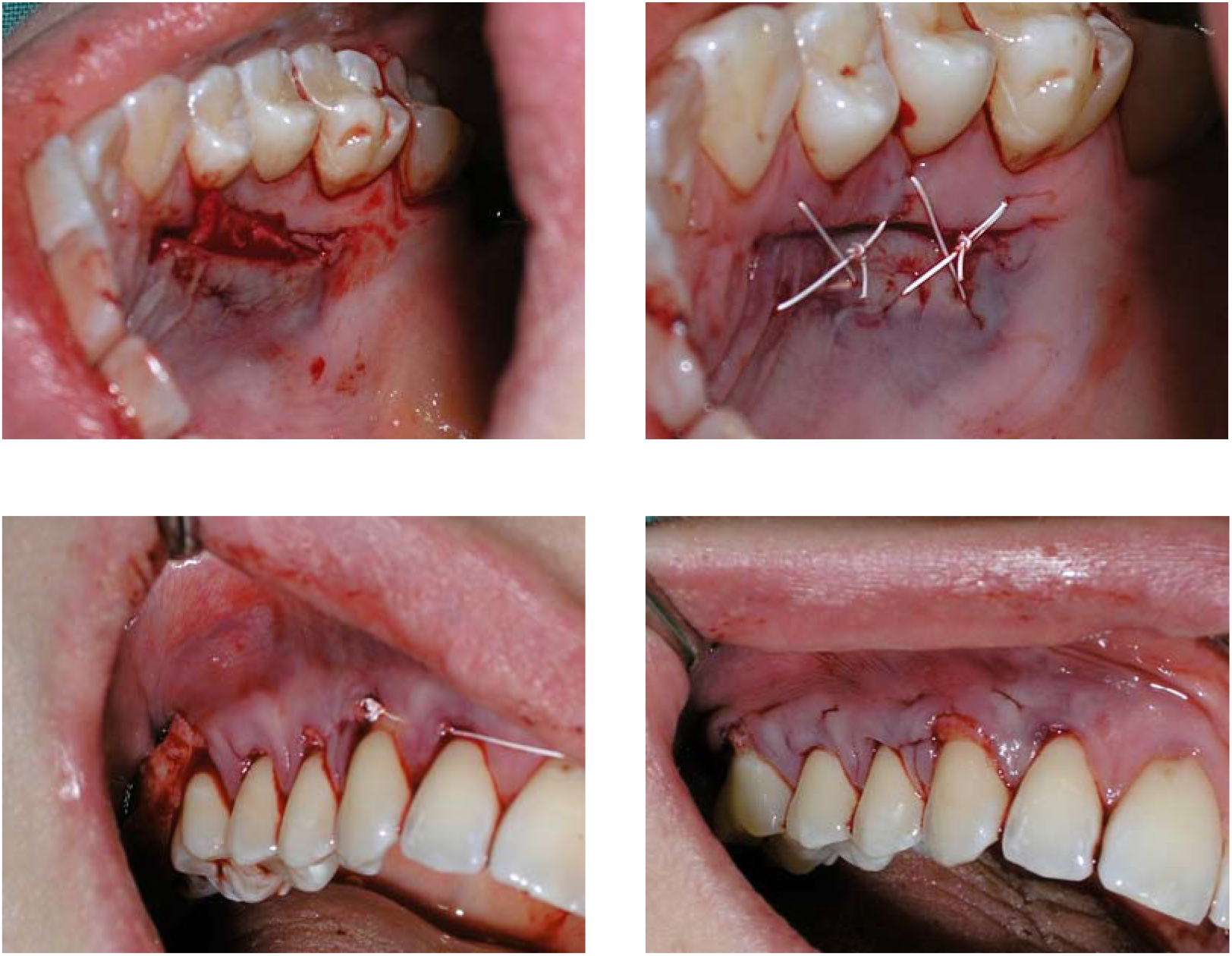
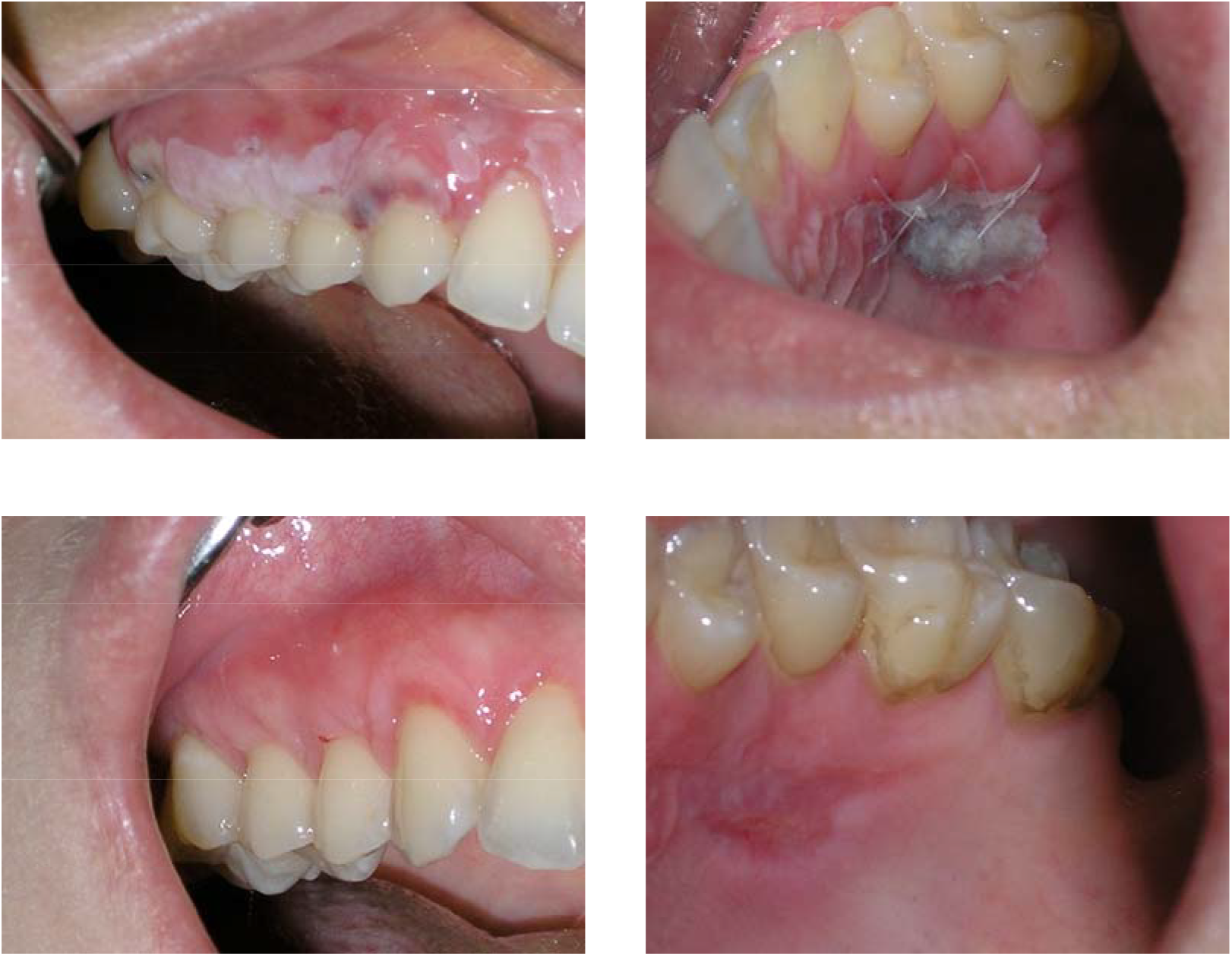
Representative case illustrating the single-incision graft-harvesting technique, tunnel surgery, early postoperative healing, and the final clinical result.

### 3.3 Long-term marginal stability

At the final available follow-up, no treated site exhibited a clinically relevant relapse of ≥1 mm after initial healing. Marginal soft-tissue levels therefore remained stable according to the prespecified clinical definition throughout the documented observation period, including cases followed for up to 16 years.

### 3.4 Site-level observations

Favorable treatment responses were documented in both anterior and posterior regions and in both dental arches. Multiple treated sites within individual patients generally showed comparable response patterns. The overall CRC rate of 83.4% falls within the range reported in controlled investigations of tunnel-based root-coverage procedures.

## 4. Discussion

This retrospective cohort provides long-term real-world evidence on tunnel-based root coverage with SCTG. Across 710 recession sites, mean recession reduction was 2.72 mm and CRC was achieved at 83.4% of sites. Most importantly, the available longitudinal documentation did not identify a clinically relevant relapse of ≥1 mm after initial healing, with individual observations extending to 16 years.

The observed CRC rate is consistent with the favorable outcomes reported in systematic reviews and randomized clinical trials of tunnel procedures. The principal contribution of the present cohort is not the demonstration of short-term efficacy, which is already well established, but the documentation of sustained clinical performance over substantially longer periods in routine practice.

Several biological and technical mechanisms may contribute to this durability. The tunnel approach preserves the interdental papillae and avoids vertical releasing incisions, thereby maintaining a broad vascular supply to the mobilized flap and the graft. The autogenous SCTG increases soft-tissue thickness and may convert a vulnerable thin phenotype into a more stable tissue complex. Long-term analyses of periodontal plastic-surgery cohorts have identified phenotype modification and tissue-thickness gain as important predictors of gingival-margin stability.

Supportive periodontal care is another plausible contributor. Patients in this cohort were maintained on individualized recall schedules, generally at 3- to 12-month intervals. Long-term root-coverage outcomes should therefore be interpreted as the result of both the surgical intervention and continued control of plaque, inflammation, and traumatic oral-hygiene habits.

The use of EMD in selected cases introduces clinical heterogeneity. Because the present analysis was not designed as a randomized comparison of SCTG alone versus SCTG plus EMD, no causal inference regarding the adjunctive effect of EMD can be made. Likewise, the absence of a parallel control group prevents direct comparison with coronally advanced flap procedures, collagen matrices, or acellular dermal substitutes.

The study has additional limitations. Its retrospective design limits control over confounding and the standardization of follow-up intervals. Site-level analysis creates potential within-patient dependence, although clustering was addressed statistically. Standardized three-dimensional volumetric STL measurements were not available across the full observation period, the Root Coverage Esthetic Score was not recorded consistently, and patient-reported outcome measures were incomplete. These limitations restrict comparisons with contemporary digitally documented randomized trials.

Conversely, the cohort has clinically relevant strengths. It includes a large number of recession sites, reflects treatment by a consistent operator in routine practice, and contains observations extending far beyond the follow-up of most root-coverage trials. The findings therefore complement controlled studies by providing information on the durability of phenotype-enhancing periodontal plastic surgery in a real-world setting.

Future prospective studies should combine standardized recession measurements with digital three-dimensional volumetric analysis, calibrated esthetic scoring, patient-reported outcomes, and predefined long-term recall points. Stratified analyses according to recession type, tooth region, smoking status, periodontal phenotype, EMD use, and maintenance adherence would further clarify which factors determine very long-term stability.

## 5. Conclusions

Within the limitations of this retrospective longitudinal cohort study, the tunnel technique combined with subepithelial connective tissue grafting was associated with predictable root coverage and durable long-term marginal stability. Across 710 recession sites, mean recession reduction was 2.72 mm and complete root coverage was achieved at 83.4% of sites. No clinically relevant relapse of ≥1 mm after initial healing was documented at the final available follow-up, with observations extending to 16 years. These findings support minimally invasive tunnel surgery with autogenous SCTG as a durable phenotype-enhancing approach for the treatment of localized and multiple gingival recessions.

## Data Availability

All data produced in the present work are contained in the manuscript.

## 6. Ethics Approval and Consent to Participate

The study was conducted in accordance with the principles of the Declaration of Helsinki. All patients provided written informed consent for treatment and for the pseudonymized scientific use of their clinical data.

In accordance with the Federal Law Gazette Part I 2024, issued in Bonn on 26 March 2024, the patient management system used for the study provides for the use of health data for research purposes oriented towards the common good and for the data-based further development of the health care system (Health Data Use Act – GDNG), Act on the Improved Use of Health Data, a pseudonymized use of patient data, signed at the time of the recording of the patients.

## 7. Data Availability

The data underlying the findings of this study are available from the corresponding author upon reasonable request, subject to applicable data-protection requirements and restrictions relating to patient confidentiality.

## 10. Author Contributions

Jennifer Schmücker: clinical investigation, surgical treatment, data acquisition, data curation, methodology, and manuscript review and editing.

Emily Speer: manuscript review and editing.

Marco Alexander Vukovic: manuscript review and editing.

Wolf-D. Grimm: conceptualization, methodology, scientific interpretation, supervision, manuscript drafting, and manuscript review and editing.

All authors read and approved of the final manuscript.

